# Effect of the Neutralizing SARS-CoV-2 Antibody Sotrovimab in Preventing Progression of COVID-19: A Randomized Clinical Trial

**DOI:** 10.1101/2021.11.03.21265533

**Authors:** Anil Gupta, Yaneicy Gonzalez-Rojas, Erick Juarez, Manuel Crespo Casal, Jaynier Moya, Diego Rodrigues Falci, Elias Sarkis, Joel Solis, Hanzhe Zheng, Nicola Scott, Andrea L. Cathcart, Sergio Parra, Jennifer E. Sager, Daren Austin, Amanda Peppercorn, Elizabeth Alexander, Wendy W. Yeh, Cynthia Brinson, Melissa Aldinger, Adrienne E. Shapiro

## Abstract

**Importance:** Older patients and those with underlying comorbidities infected with SARS-CoV-2 may be at increased risk of hospitalization and death from COVID-19. Sotrovimab is a neutralizing antibody designed for treatment of high-risk patients to prevent COVID-19 progression.

**Objective:** To evaluate the efficacy and safety of sotrovimab in preventing progression of mild to moderate COVID-19 to severe disease.

**Design:** Randomized, double-blind, multicenter, placebo-controlled, phase 3 study.

**Setting:** 57 centers in 5 countries.

**Participants:** Nonhospitalized patients with symptomatic, mild to moderate COVID-19 and at least 1 risk factor for disease progression.

**Intervention:** Patients were randomized (1:1) to an intravenous infusion of sotrovimab 500 mg or placebo.

**Main Outcomes and Measures:** The primary efficacy outcome was the proportion of patients with COVID-19 progression, defined as all-cause hospitalization longer than 24 hours for acute illness management or death through day 29. Key secondary outcomes included the proportion of patients with COVID-19 progression, defined as emergency room visit, hospitalization of any duration, or death, and proportion of patients developing severe/critical respiratory COVID-19 requiring supplemental oxygen.

**Results:** Among 1057 patients randomized (sotrovimab, 528; placebo, 529), all-cause hospitalization longer than 24 hours or death was significantly reduced with sotrovimab (6/528 [1%]) vs placebo (30/529 [6%]) by 79% (95% CI, 50% to 91%; *P*<.001). Secondary outcome results further demonstrated the effect of sotrovimab in reducing emergency room visits, hospitalization of any duration, or death, which was reduced by 66% (95% CI, 37% to 81%; *P*<.001), and severe/critical respiratory COVID-19, which was reduced by 74% (95% CI, 41% to 88%; *P*=.002). No patients receiving sotrovimab required high-flow oxygen, oxygen via nonrebreather mask, or mechanical ventilation compared with 14 patients receiving placebo. The proportion of patients reporting adverse events was similar between treatment groups; sotrovimab was well tolerated, and no safety concerns were identified.

**Conclusions and Relevance:** Among nonhospitalized patients with mild to moderate COVID-19, a single 500-mg intravenous dose of sotrovimab prevented progression of COVID-19, with a reduction in hospitalization and need for supplemental oxygen. Sotrovimab is a well-tolerated, effective treatment option for patients at high risk for severe morbidity and mortality from COVID-19.

**Trial Registration:** ClinicalTrials.gov Identifier: <u>NCT04545060</u>

## Introduction

Over 4.8 million people worldwide have died from COVID-19.^1,2^ The most common serious manifestations of COVID-19 are respiratory failure and acute respiratory distress syndrome, but diverse effects have been observed in other organ systems.^3^ Patient characteristics associated with a greater risk of severe COVID-19 include older age, obesity, diabetes mellitus, chronic obstructive pulmonary disease, and chronic kidney disease.^4-9^

Since the onset of the global pandemic in March 2020, mutations in the spike gene of SARS-CoV-2 have resulted in the global spread of variants of concern that may increase transmissibility and disease severity while decreasing response to preventative measures and treatment options.^10-13^ The B.1.617.2 (Delta) variant that originated in India in late 2020 has emerged as the leading variant of concern to date, with increased transmissibility and immune evasion, including vaccine breakthrough infections.^14^ In the United States, current treatment guidelines for outpatients with mild to moderate COVID-19 who are at high risk for clinical progression recommend either casirivimab plus imdevimab or sotrovimab regardless of region.^15^

Given the evolution of SARS-CoV-2 variants, limited worldwide vaccine availability, reduced vaccine efficacy in certain immunocompromised populations, and vaccine hesitancy,^10,16,17^ there is a need for effective therapies that provide a high barrier against viral escape and will provide enduring coverage as the virus continues to evolve.^14^

Sotrovimab (VIR-7831) is an Fc-engineered human monoclonal antibody developed from a parental antibody isolated from a survivor of the SARS outbreak in 2003 and contains the “LS” modification to enhance half-life and respiratory mucosal delivery.^18-22^ In contrast to other monoclonal antibodies,^23-26^ sotrovimab targets a highly conserved epitope in the SARS-CoV-2 spike protein at a region that does not compete with angiotensin-converting enzyme 2 binding.^19^ In addition to neutralizing SARS-CoV-2, sotrovimab has demonstrated potent effector functions in vitro that may contribute to immune-mediated viral clearance.^18,19^ Notably, data also suggest that sotrovimab may prevent cell-cell fusion (ie, syncytia formation), unlike other receptor binding domain–targeting antibodies.^27^

The <u>CO</u>vid-19 <u>M</u>onoclonal antibody <u>E</u>fficacy <u>T</u>rial-Intent to <u>C</u>are <u>E</u>arly (COMET-ICE) trial evaluated the efficacy and safety of sotrovimab administered intravenously in high-risk patients with mild to moderate COVID-19. Results from a preplanned interim analysis of data including 583 patients were recently published.^28^ Enrollment was stopped early due to overwhelming efficacy at the time of the interim analysis, with an 85% reduction in all-cause hospitalization for over 24 hours or death in patients treated with sotrovimab compared with those who received placebo. The current report presents the full results of COMET-ICE through the primary endpoint at day 29 for the intent-to-treat (ITT) population of high-risk, ambulatory patients with mild to moderate COVID-19.

## Methods

### Study Design

The early treatment of mild to moderate COVID-19 with sotrovimab was assessed in this phase 3, randomized, double-blind, placebo-controlled, multicenter study. There were 57 participating centers (United States, 45; Brazil, 6; Spain, 3; Canada, 2; Peru, 1). The study was conducted in accordance with the principles of the Declaration of Helsinki and Council for International Organizations of Medical Sciences International Ethical Guidelines, applicable International Council for Harmonisation Good Clinical Practice guidelines, and applicable laws and regulations. Written informed consent was provided by all patients prior to study entry; patients did not receive a stipend.

### Patients

Eligible patients were 18 years of age or older, tested positive for SARS-CoV-2 by reverse-transcriptase polymerase chain reaction (RT-PCR) or antigen test, and had symptom onset within the prior 5 days. The study population represented patients at high risk for COVID-19 progression to hospitalization or death. As such, patients were required to have at least 1 of the following risk factors: age 55 years or older, diabetes requiring medication, obesity (body mass index >30 kg/m^2^), chronic kidney disease (estimated glomerular filtration rate <60 mL/min/1.73 m^2^),^29^ congestive heart failure (New York Heart Association class II or higher), chronic obstructive pulmonary disease, or moderate to severe asthma.^30^ Patients were excluded if they were hospitalized or if they had signs/symptoms of severe COVID-19 (shortness of breath at rest, oxygen saturation <94%, or requiring supplemental oxygen).

### Randomization and Intervention

Eligibility screening was performed within 24 hours before study drug administration. Using an interactive web response system, eligible patients were randomized 1:1 to receive a single intravenous infusion of sotrovimab 500 mg or an equal volume of saline placebo over 1 hour on day 1. Patients were observed for approximately 2 hours after infusion. Patients were stratified by age (≤70 vs >70 years), duration of COVID-19 symptoms (≤3 vs 4-5 days), and region (North America vs South America vs Europe).

### Primary and Secondary Outcomes

The primary outcome was the proportion of patients with all-cause hospitalization for more than 24 hours or death through day 29. To capture clinical events that may not have required prolonged hospitalization but were potentially clinically relevant, a secondary outcome of the proportion of patients with all-cause emergency room (ER) visits, hospitalizations of any duration for acute illness management, or death through day 29 was measured. Additional prespecified clinical secondary outcomes were the proportion of patients with progression of COVID-19 to severe/critical disease (defined as requirement for supplemental oxygen [severe disease] or mechanical ventilation [critical disease]) through day 29, and all-cause mortality at day 29. Symptom severity and duration were measured by the COVIDLJ19–adapted version of the inFLUenza Patient-Reported Outcome (FLU-PRO) Plus tool and assessed as mean change in total score from baseline through day 7 (see **Supplement 1** for more information on the FLU-PRO Plus).^31^ Changes from baseline to day 8 in viral load in nasal secretions were determined by quantitative RT-PCR.

### Exploratory Outcomes

Prespecified exploratory outcome measures included total hospital length of stay, total intensive care unit (ICU) length of stay, and total number of ventilator days from randomization through day 29.

### Safety Outcomes

Adverse events (AEs) and serious AEs, including all hospitalizations and deaths, regardless of relationship to COVID-19, were assessed. AEs of special interest were defined as infusion-related reactions (including hypersensitivity reactions) and the potential for antibody-dependent enhancement.

### Sample Size

A sample size of 1360 patients (680 per treatment group) was determined to provide approximately 90% power to detect a 37.5% relative efficacy in reducing COVID-19 progression through day 29 at the overall 2-sided 5% significance level. The assumed progression of COVID-19 rates was 16% in the placebo group and 10% in the sotrovimab group.

### Statistical Analysis

The ITT population included all randomized patients, irrespective of treatment received. The safety population included all patients who received study treatment. The virology population was a subset of the ITT population including patients with a central laboratory–confirmed quantifiable baseline nasopharyngeal swab.

The study utilized a group sequential design with 2 interim analyses to assess both futility due to lack of efficacy and overwhelming efficacy. A Lan-DeMets^32^ alpha-spending function to control the type I error for the primary endpoint was used, with a Pocock analog rule for futility and a Hwang-Shih-DeCani (γ=1) analog rule for efficacy.^33^ The primary endpoint was analyzed in the ITT population using a Poisson regression model with robust sandwich estimators adjusting for the duration of symptoms, age, and sex. Missing data were imputed under a missing at random assumption using a multiple imputation model. Secondary outcomes were formally analyzed in the ITT population (except the viral load outcome, which was assessed in the virology population) at the final day 29 analysis using a 2-sided alpha level of 5%. Statistical testing of secondary outcomes was adjusted for multiplicity using a hierarchy in which the full alpha (*P*≤.05) was transferred down between each outcome measure. The order of hierarchical testing of secondary outcomes was COVID-19 progression (all-cause ER visits, hospitalizations of any duration, or deaths), change in viral load, severe/critical respiratory COVID-19 requiring supplemental oxygen, change in FLU-PRO Plus total score, and all-cause mortality. The proportion of patients with all-cause ER visits, hospitalizations of any duration, or death and the proportion of patients with severe/critical respiratory COVID-19 were analyzed in a similar manner as the primary endpoint. Mean change in FLU-PRO Plus was analyzed as area under the curve (AUC) through day 7 using analysis of covariance adjusted for baseline value, age group, time to symptom onset, sex, and region. Mean change in log_10_-transformed nasal viral load was analyzed using a mixed model for repeated measures model adjusted for baseline value, baseline value by visit, age, duration of symptoms, and sex.

Statistical analyses were performed using SAS version 9.4.

## Results

### Patient Demographics and Clinical Characteristics

Of 1351 patients screened from August 2020 through March 2021, 1057 were randomly assigned to sotrovimab (n=528) or placebo (n=529), comprising the ITT population (**Figure 1**). Database lock occurred in April 2021. Eight randomized patients did not receive study drug and were not included in the safety population (n=1049). The median (range) duration of follow-up was 103 (5 to 178) and 102 (3 to 176) days for the sotrovimab and placebo group, respectively.

**Figure 1.**
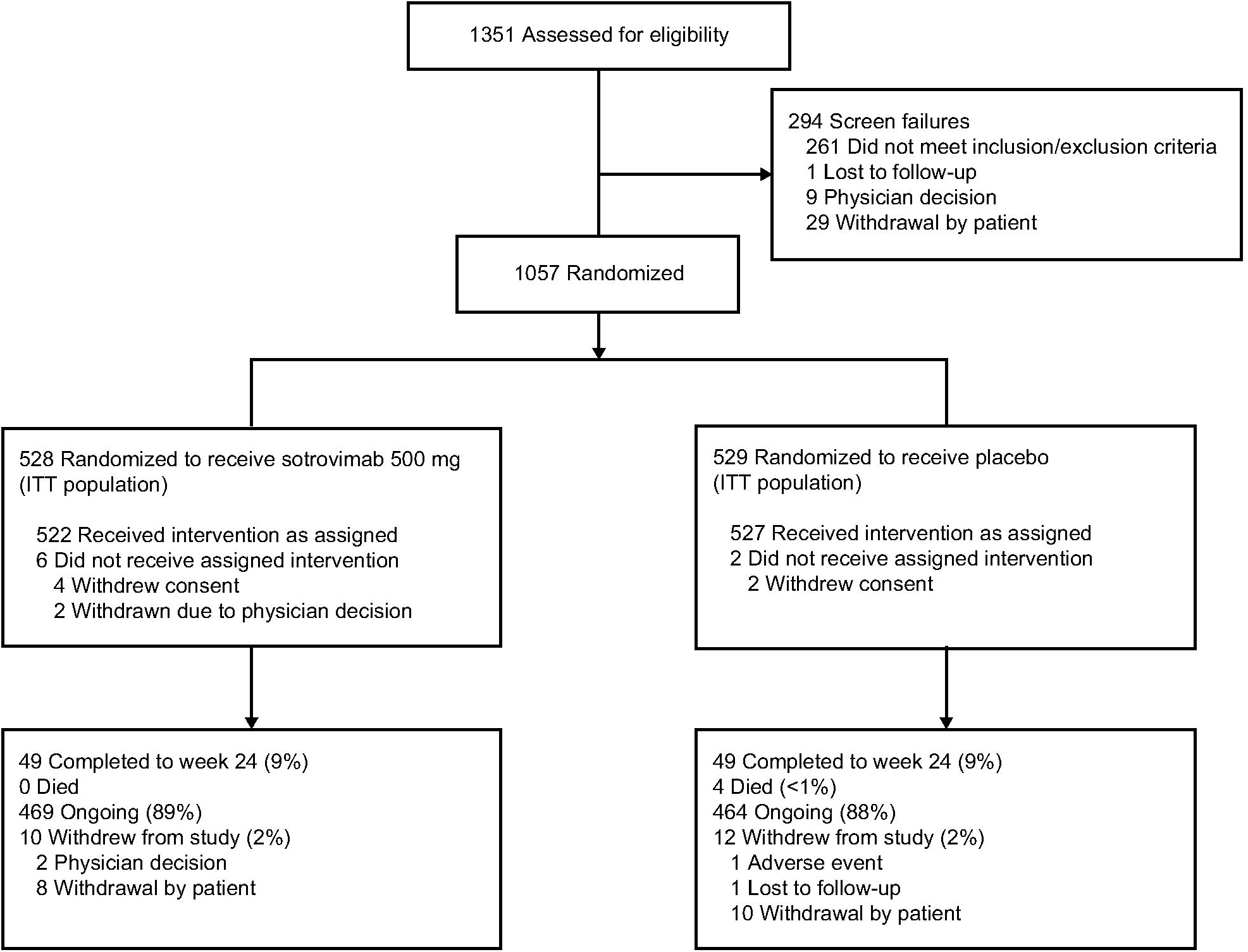
Patient Enrollment and Treatment Assignment in the COMET-ICE Trial^a^. ITT, intent-to-treat; AE, adverse event. ^a^One patient randomized to placebo received sotrovimab and is included in the sotrovimab group of the safety population but in the placebo group of the ITT population; this patient had no AEs or hospitalizations.

Baseline demographics and clinical characteristics were well balanced between treatment groups (**Table 1**). The median age was 53 years, with 20% of patients aged 65 years or older. The majority of patients (65%) were Latinx. The 4 most common predefined risk factors or comorbidities in both treatment groups at screening were obesity, age 55 years or older, diabetes requiring medication, and moderate to severe asthma. Most patients (59%) had symptoms for 3 or fewer days. Presenting symptoms were similar between treatment groups, with cough, headache, myalgia, and fatigue most common.

**Table 1.**
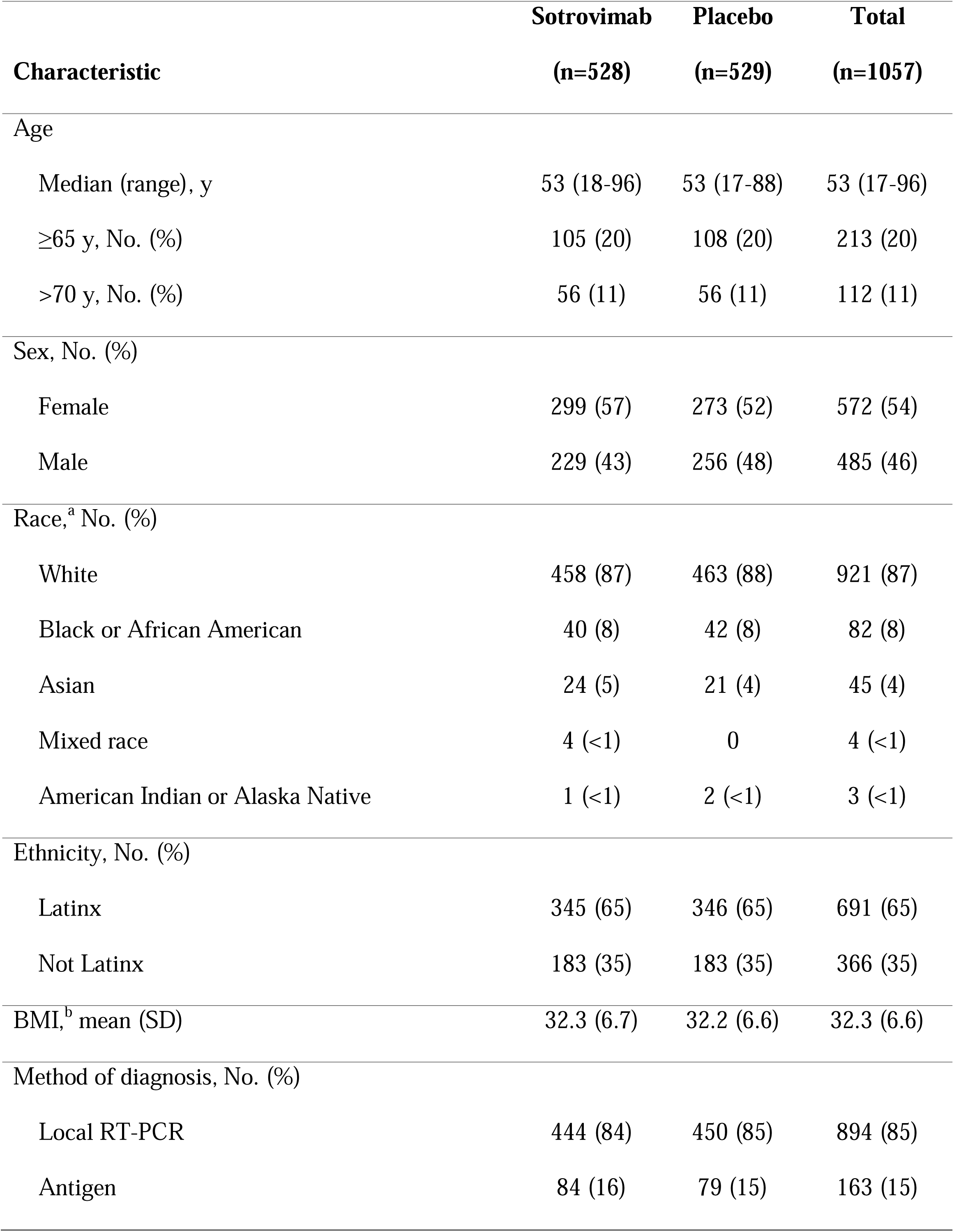
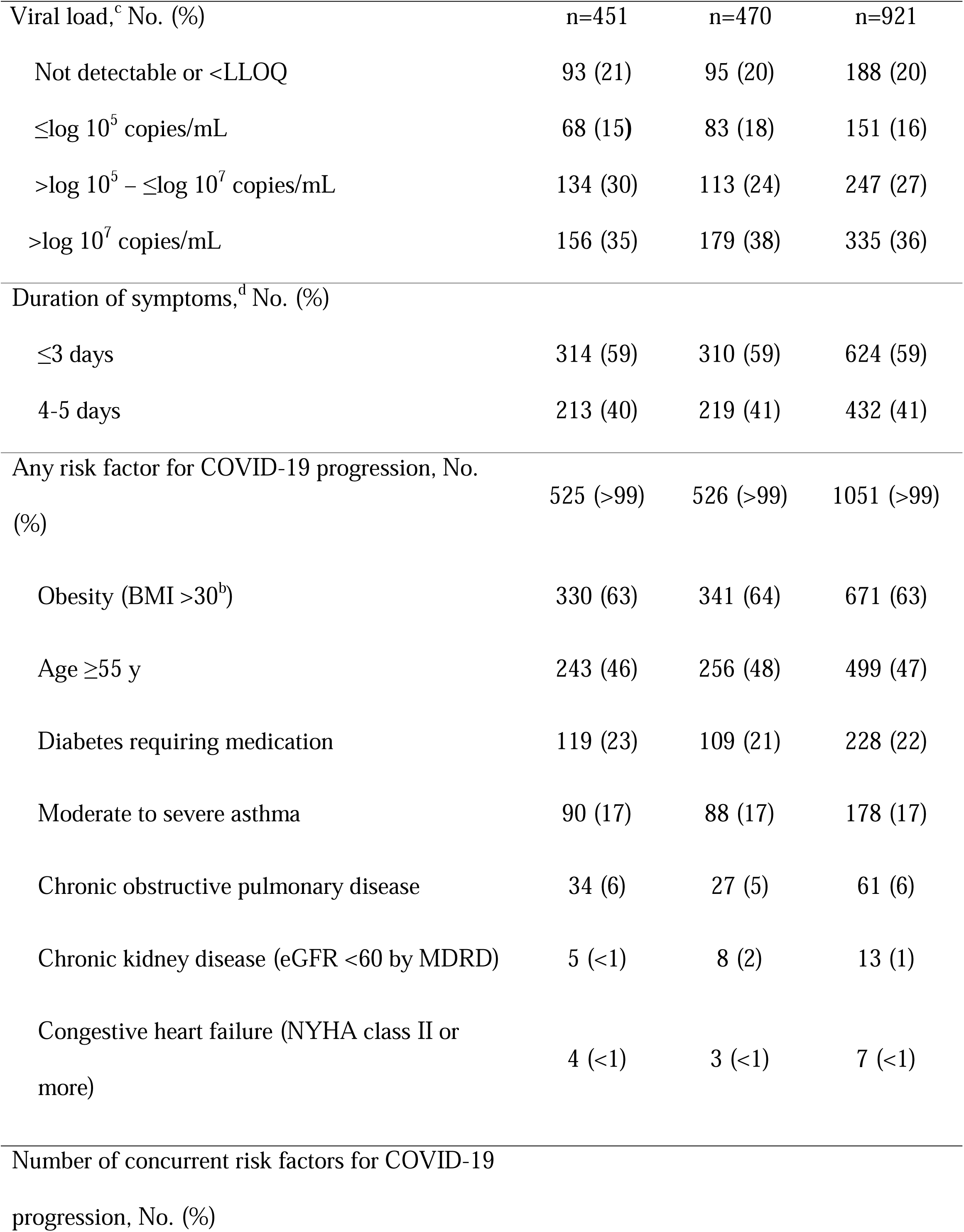
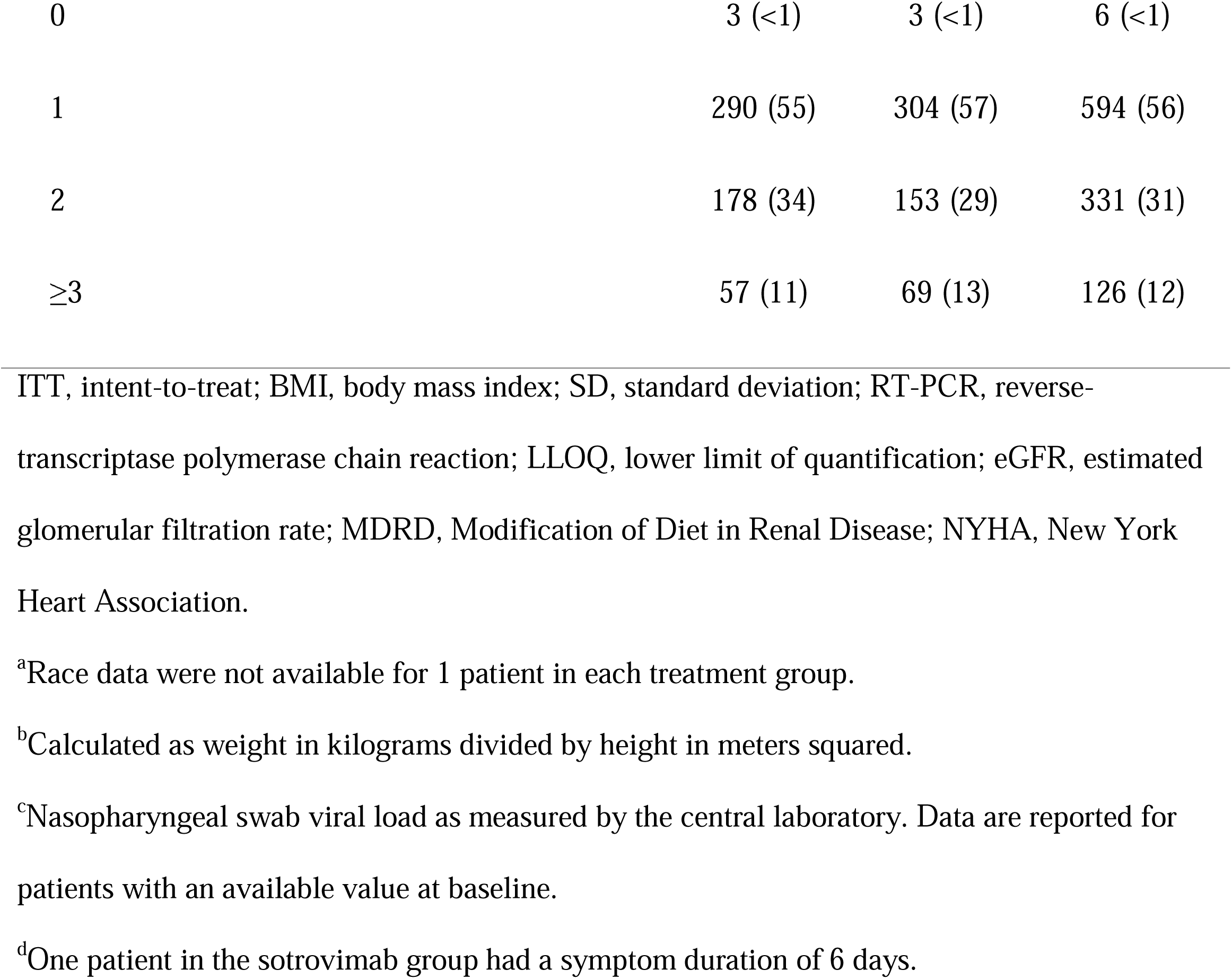
Patient Demographics and Baseline Disease Characteristics (ITT Population)

### Primary and Secondary Outcomes

Six of 528 (1%) patients treated with sotrovimab compared with 30 of 529 (6%) patients receiving placebo progressed to hospitalization (>24 hours, any cause) or death (any cause) through day 29, resulting in a statistically significant reduction of 79% (95% CI, 50% to 91%; *P*<.001; **Table 2**). In a post-hoc review, of the patients receiving sotrovimab who were hospitalized, 3 had respiratory conditions associated with COVID-19 and 3 were hospitalized for other reasons (small intestinal obstruction, non-small cell lung cancer, and diabetic foot ulcer; **eTable 1** in **Supplement 1)**.

**Table 2.**
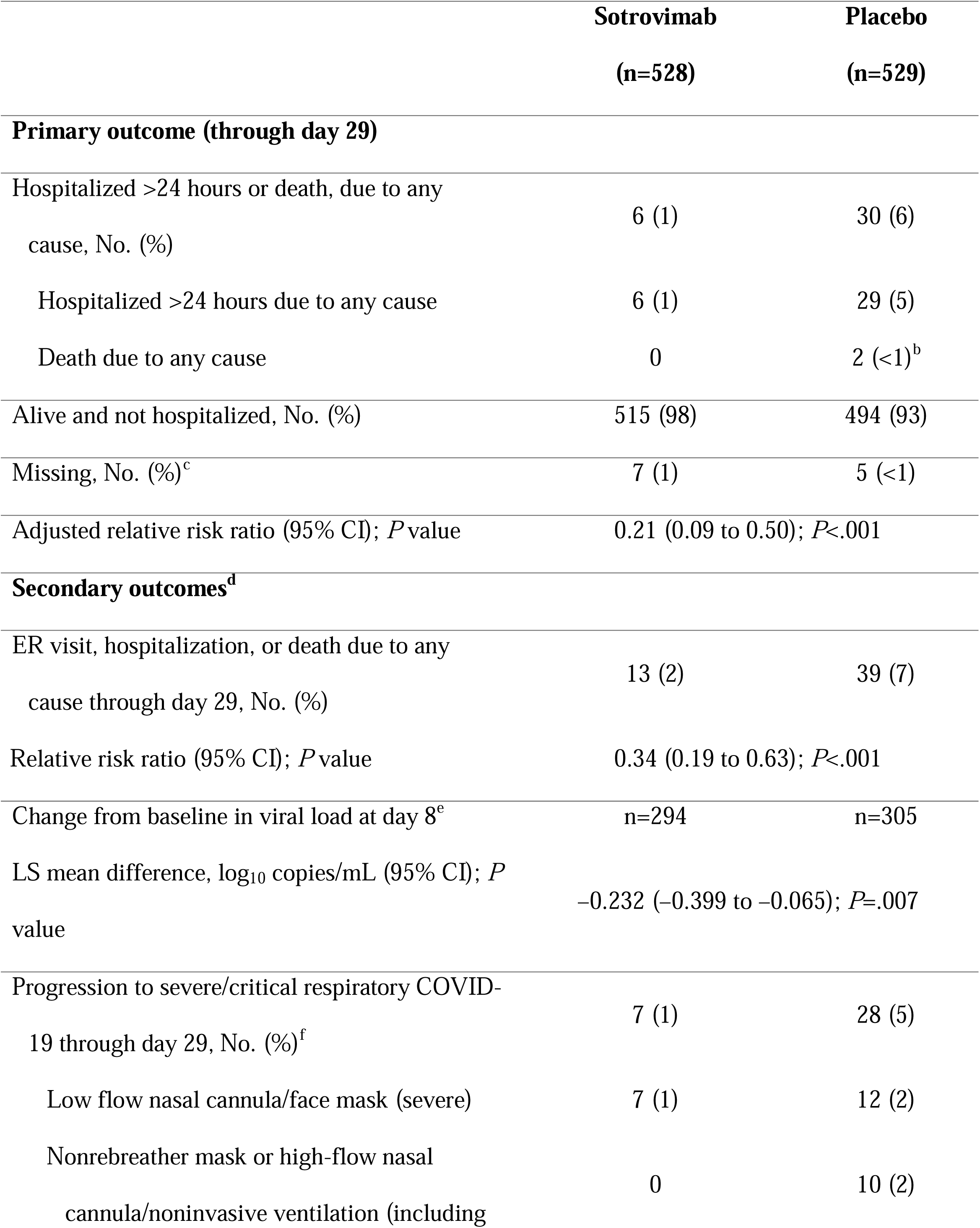
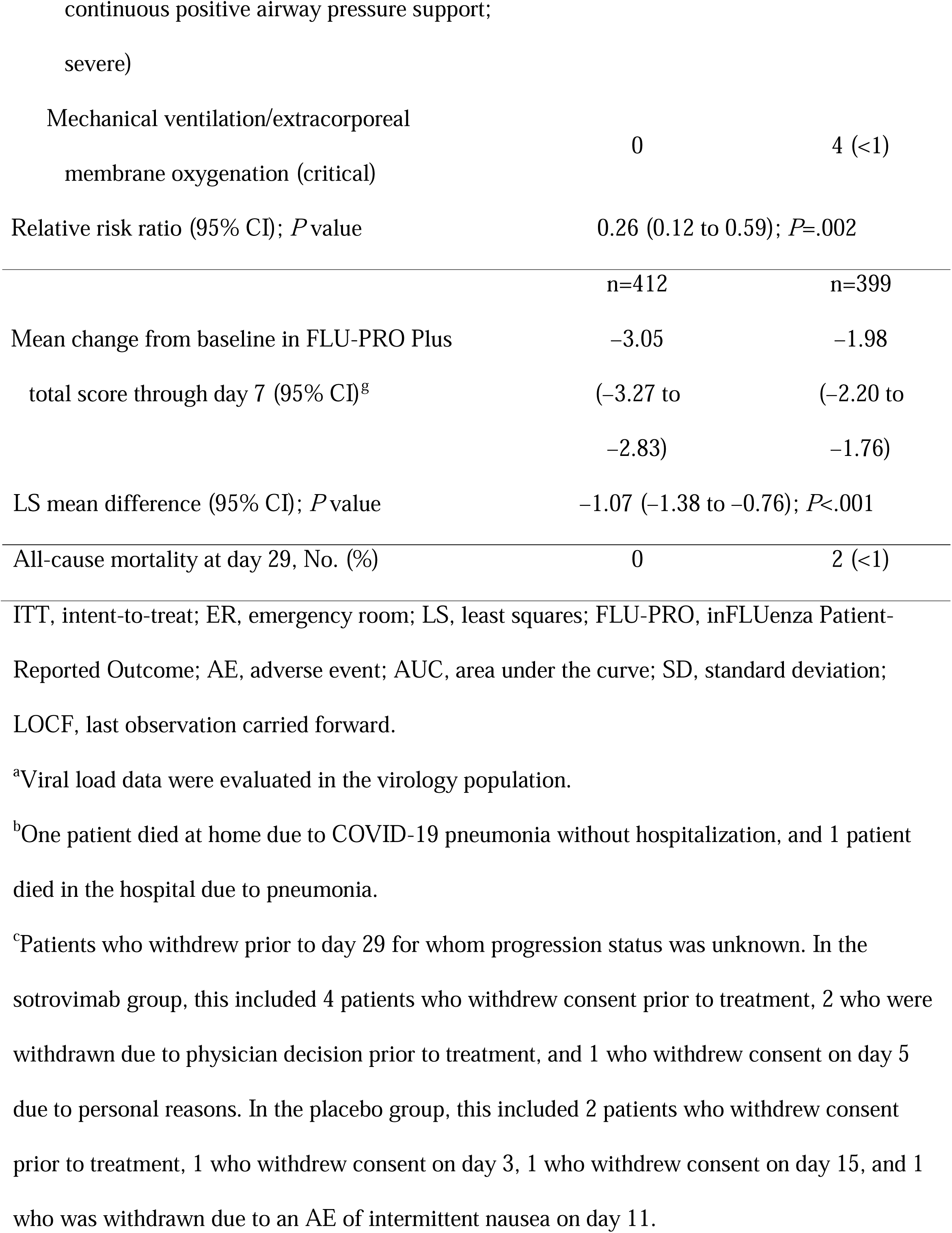
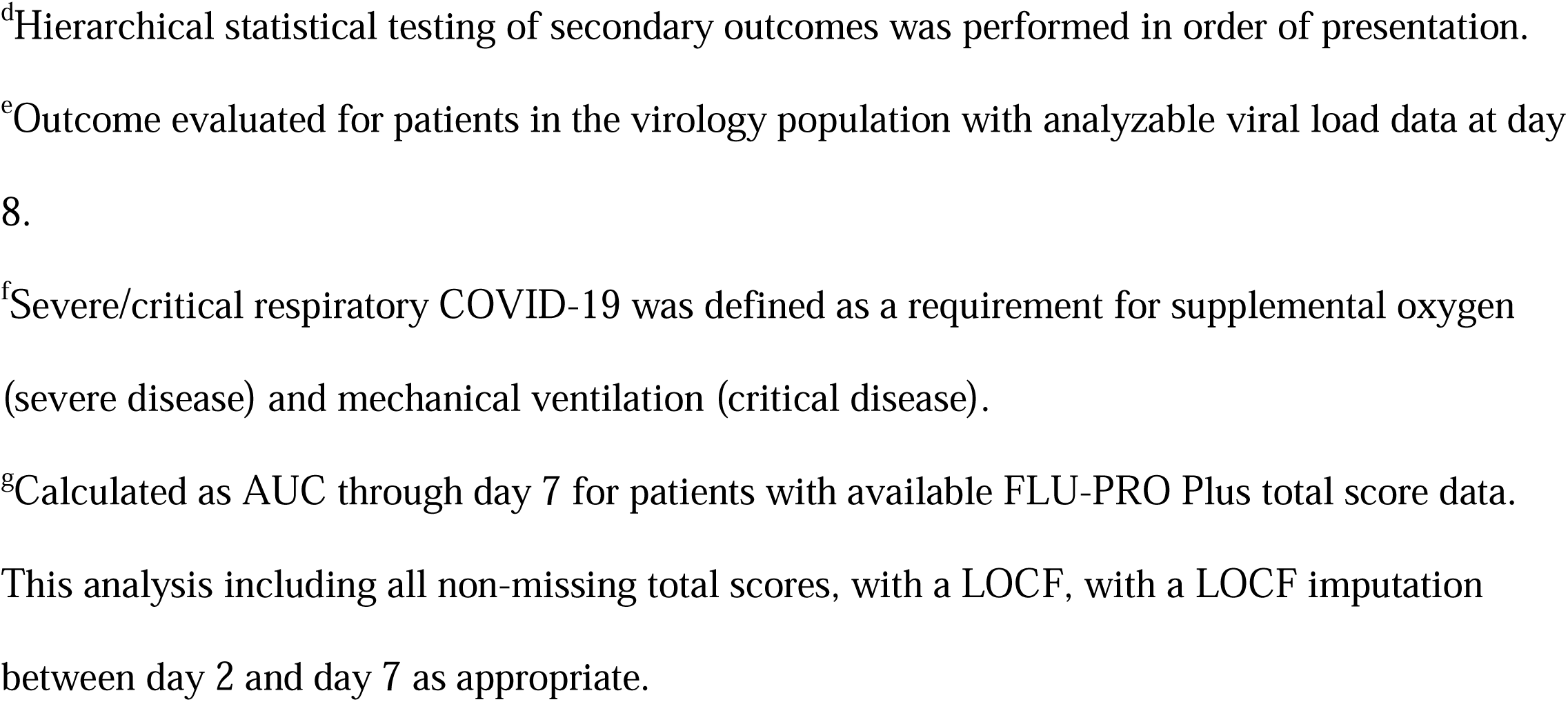
Primary and Secondary Efficacy Outcomes (ITT Population^a^)

All secondary endpoints tested in the hierarchy met statistical significance except all-cause mortality, which was not formally analyzed due to fewer than anticipated deaths (**Table 2**). The percentage of patients who progressed to all-cause ER visit, any duration of hospitalization, or death, was reduced by 66% with sotrovimab vs placebo (95% CI, 37% to 81%; *P*<.001). Similarly, sotrovimab significantly reduced progression to severe/critical respiratory COVID-19 compared with placebo (adjusted relative risk reduction: 74%; 95% CI, 41% to 88%; *P*=.002). No patients treated with sotrovimab required high-flow oxygen, oxygen via a nonrebreather mask, or mechanical ventilation. Among patients who received placebo, 10 required oxygen support (high-flow nasal cannula, nonrebreather mask, or noninvasive ventilation) and 4 required mechanical ventilation (**eFigure 1** in **Supplement 1**). By day 29, there were no deaths in the sotrovimab group and 2 deaths in the placebo group.

Among patients in the virology population (n=733), the mean decline from baseline in viral load at day 8 was significantly greater with sotrovimab vs placebo (difference: –0.232 log_10_ copies/mL; 95% CI, –0.399 to –0.065; *P*=.007; **Table 2**). Among patients in the ITT population, 57% of those receiving sotrovimab and 56% of those receiving placebo completed the FLU-PRO Plus questionnaire through day 7. Mean decreases in total score AUC from baseline to day 7 were significantly greater in the sotrovimab group compared with the placebo group (difference: –1.07; 95% CI, –1.38 to –0.76; *P*<.001).

### Exploratory Outcomes

Treatment with sotrovimab reduced the number of patients who were admitted to the hospital for more than 24 hours, for any reason, compared with placebo; among patients who required hospitalization, treatment with sotrovimab resulted in numerical reductions in the duration of hospitalization compared with placebo (**Table 3**). Additionally, no patients receiving sotrovimab required an ICU stay or mechanical ventilator support while hospitalized compared with 10 (2%) and 6 (1%) patients receiving placebo, respectively.

**Table 3.** Exploratory Outcomes Through Day 29 (ITT Population)

|  | **Sotrovimab** | **Placebo** |
| --- | --- | --- |
|  | **(n=528)** | **(n=529)** |
| Hospital length of stay, No. (%) |  |  |
| 0 days | 521 (99) | 499 (94) |
| >0 to ≤24 hours | 1 (<1) | 0 |
| 1 to ≤8 days | 3 (<1) | 19 (4) |
| 9 to ≤15 days | 2 (<1) | 3 (<1) |
| 16 to ≤22 days | 1 (<1) | 4 (<1) |
| 23 to ≤29 days | 0 | 4 (<1) |
| ICU length of stay, No. (%) |  |  |
| 0 days | 528 (100) | 519 (98) |
| 1 to ≤8 days | 0 | 2 (<1) |
| 9 to ≤15 days | 0 | 3 (<1) |
| 16 to ≤22 days | 0 | 2 (<1) |
| 23 to ≤29 days | 0 | 3 (<1) |
| Duration of ventilator use, No. (%) |  |  |
| 0 days | 528 (100) | 523 (99) |
| 1 to ≤8 days | 0 | 0 |
| 9 to ≤15 days | 0 | 2 (<1) |
| 16 to ≤22 days | 0 | 3 (<1) |
| 23 to ≤29 days | 0 | 1 (<1) |

ITT, intent-to-treat; ICU, intensive care unit.

### Safety

In the safety analysis population (n=1049), AEs were reported for 22% (114 of 523) of patients in the sotrovimab group and 23% (123 of 526) of patients in the placebo group (**Table 4**). No deaths were reported for patients receiving sotrovimab. Four deaths occurred in patients receiving placebo (2 occurred prior to day 29, and 2 occurred after day 29), with 2 classified as COVID-19 pneumonia, 1 as pneumonia, and 1 as respiratory failure. Serious AEs and grade 3 or 4 AEs were less common in patients receiving sotrovimab compared with placebo. No serious AEs were considered related to sotrovimab.

**Table 4.**
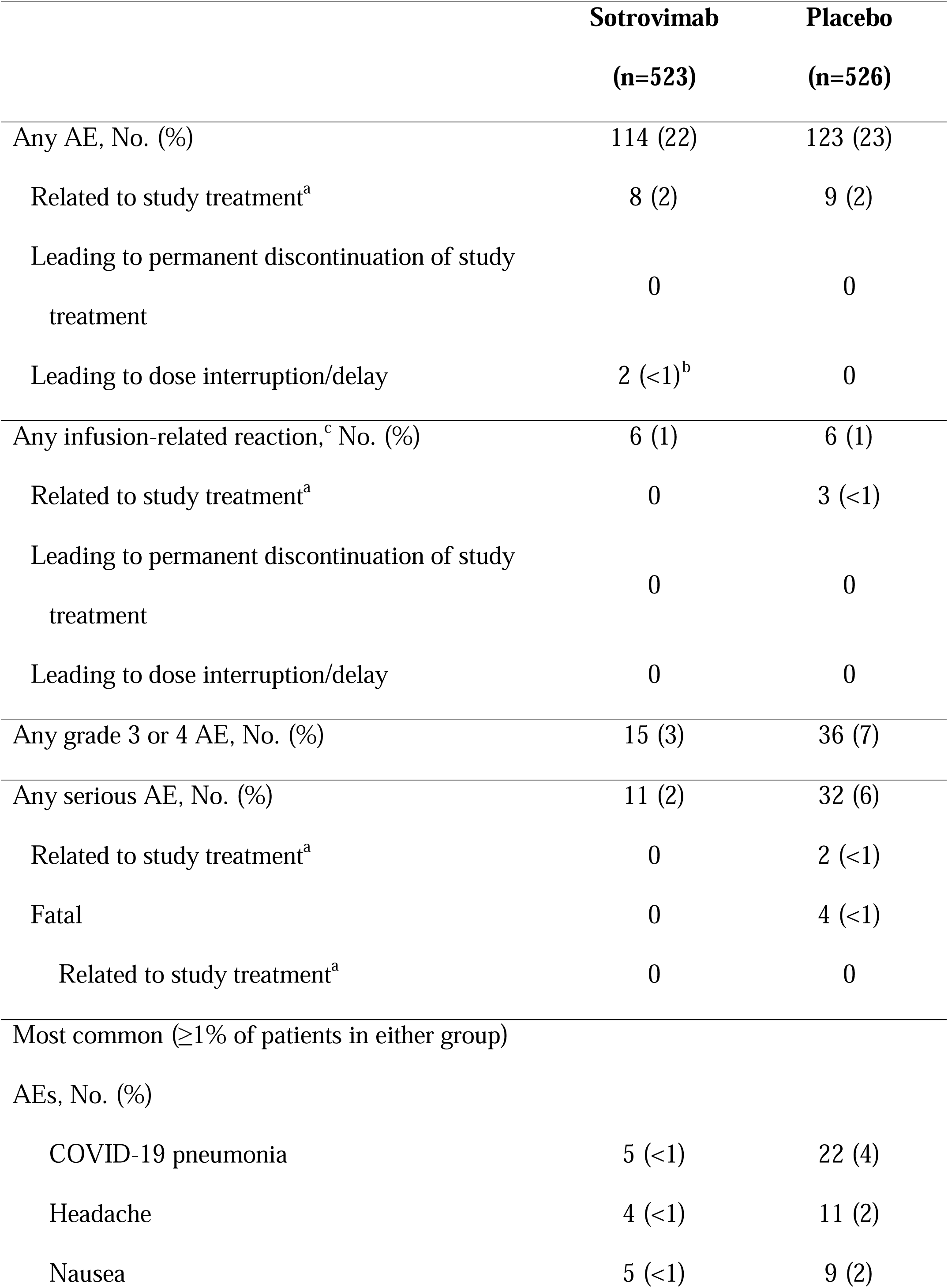
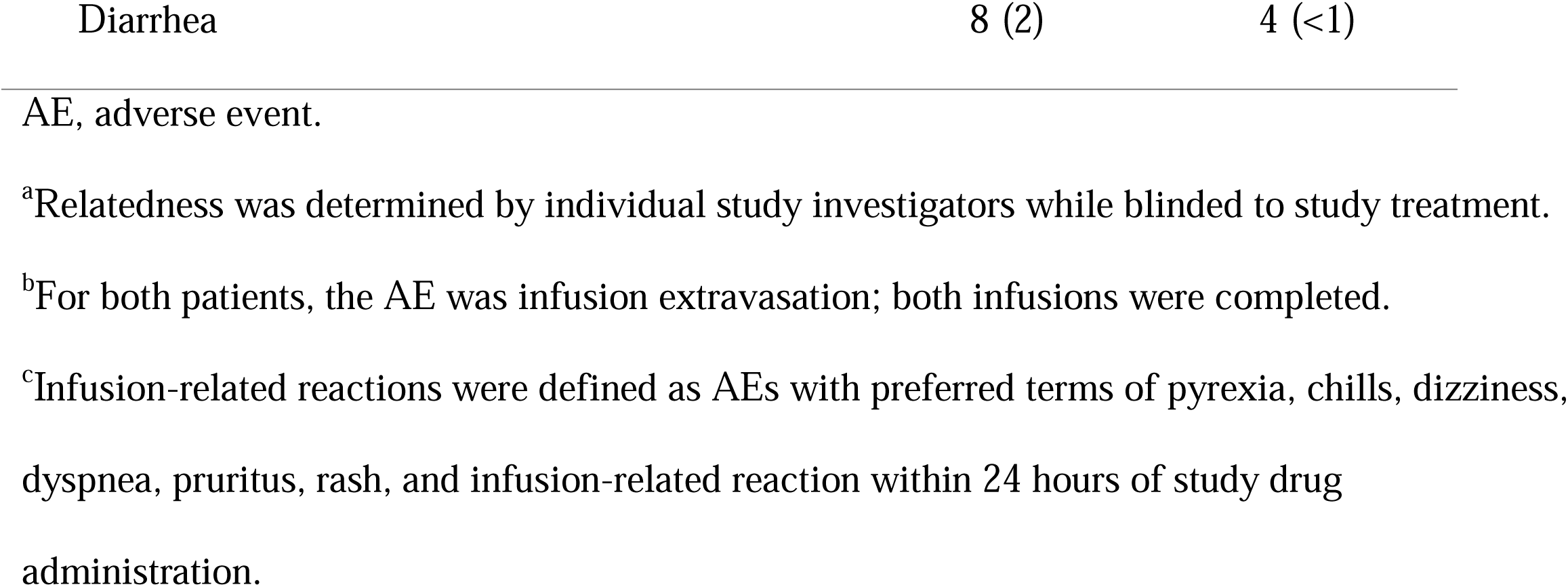
Summary of AEs (Safety Analysis Population)

|  | **Sotrovimab** | **Placebo** |
| --- | --- | --- |
|  | **(n=523)** | **(n=526)** |
| Any AE, No. (%) | 114 (22) | 123 (23) |
| Related to study treatment<sup>a</sup> | 8 (2) | 9 (2) |
| Leading to permanent discontinuation of study treatment | 0 | 0 |
| Leading to dose interruption/delay | 2 (<1)<sup>b</sup> | 0 |
| Any infusion-related reaction,<sup>c</sup> No. (%) | 6 (1) | 6 (1) |
| Related to study treatment<sup>a</sup> | 0 | 3 (<1) |
| Leading to permanent discontinuation of study treatment | 0 | 0 |
| Leading to dose interruption/delay | 0 | 0 |
| Any grade 3 or 4 AE, No. (%) | 15 (3) | 36 (7) |
| Any serious AE, No. (%) | 11 (2) | 32 (6) |
| Related to study treatment<sup>a</sup> | 0 | 2 (<1) |
| Fatal | 0 | 4 (<1) |
| Related to study treatment<sup>a</sup> | 0 | 0 |
| Most common (≥1% of patients in either group) |  |  |
| AEs, No. (%) |  |  |
| COVID-19 pneumonia | 5 (<1) | 22 (4) |
| Headache | 4 (<1) | 11 (2) |
| Nausea | 5 (<1) | 9 (2) |

AE, adverse event.
<sup>a</sup>Relatedness was determined by individual study investigators while blinded to study treatment.
<sup>b</sup>For both patients, the AE was infusion extravasation; both infusions were completed.
<sup>c</sup>Infusion-related reactions were defined as AEs with preferred terms of pyrexia, chills, dizziness, dyspnea, pruritus, rash, and infusion-related reaction within 24 hours of study drug administration.

AEs occurring in more than 1% of patients in either treatment group were more frequent in the placebo group vs the sotrovimab group except diarrhea, which occurred in 8 (2%) patients receiving sotrovimab and 4 (<1%) patients receiving placebo (**Table 4**). Of the 8 sotrovimab-treated patients who reported diarrhea, all had grade 1 or 2 events, and diarrhea resolved for all but 1 patient at the data cutoff date for the day 29 analysis.

The proportion of patients with systemic infusion-related reactions was similar in each treatment group (**Table 4**), and all reactions were grade 1 or 2 and clinically manageable. Results did not suggest antibody-dependent enhancement with sotrovimab, as worsening of disease with sotrovimab vs placebo was not observed.^34^ Changes in laboratory parameters and vital signs were consistent with underlying disease and similar in both treatment groups.

## Discussion

Despite widespread vaccination campaigns in many countries, a significant proportion of the global population remains unvaccinated, has vaccine hesitancy, and/or may be immunocompromised and thus is at risk for COVID-19.^1,16,17,35,36^ Therefore, a multiprong approach, including treatment and disease prevention, will be necessary to reduce the morbidity and mortality from the current COVID-19 pandemic. The results of the COMET-ICE trial demonstrate that a single 500-mg intravenous dose of sotrovimab resulted in a clinically and statistically significant reduction in all-cause hospitalization and/or death in high-risk patients with symptomatic, mild to moderate COVID-19. Sotrovimab was associated with a 79% reduction in the proportion of patients who progressed to hospitalization, for any cause, or died through day 29 compared with placebo. As some of the hospitalizations that met the primary endpoint appeared potentially unrelated to COVID-19 disease, a post hoc review of all safety narratives was conducted. Of the 6 hospitalizations in the sotrovimab group, 3 patients were hospitalized for events potentially unrelated to COVID-19. Thirty patients in the placebo group met progression criteria for the primary endpoint, all of which were potentially related to COVID-19. These results are consistent in magnitude with the profound efficacy observed at the interim analysis of this trial.^28^ Sotrovimab was well tolerated, and no safety concerns were identified.

In this final ITT analysis, hierarchical statistical testing was performed on the 5 key secondary endpoints. All were found to be statistically significant except all-cause mortality through day 29 due to a lower number of anticipated deaths that precluded formal analysis (**Table 2**). Statistical testing of these secondary endpoints supports the findings previously published for the 3 clinical key secondary endpoints.^28^ Specifically, no patients treated with sotrovimab required high-flow oxygen, oxygen via a nonrebreather mask, or mechanical ventilation through day 29.

Importantly, among those who were hospitalized, no patients who received sotrovimab required admission to the ICU compared with 9 patients who received placebo, suggesting that sotrovimab prevents more severe complications of COVID-19 in addition to preventing the need for hospitalization itself.

While sotrovimab also significantly reduced viral load at day 8 (consistent with the drug’s mode of action comprising virus neutralization and Fc-mediated effector function), the magnitude of these reductions was modest despite sotrovimab’s profound clinical impact. These data suggest that nasopharyngeal viral load changes alone may not be a strong predictor of clinical disease course with sotrovimab treatment. This finding is consistent with the lack of evidence indicating that antiviral activity in the lung can be accurately measured via nasopharygeal RT-PCR, due to the anatomic site and the fact that viral RNA may persist in the absence of replication-competent virus.^37,38^

From the patient perspective, improvements in symptoms of COVID-19 were reported, with mean decreases in FLU-PRO Plus total score significantly greater in the sotrovimab group compared with the placebo group through day 7. Together, the nonclinical key secondary outcome results support the primary endpoint of hospitalization and death and demonstrate treatment benefits of sotrovimab that are relevant to patients.

This is one of the first prospective trials of a monoclonal antibody targeting SARS-CoV-2 that was powered to evaluate a pandemic-relevant clinical outcome, progression of COVID-19 in high-risk patients. Nearly half of all patients had 2 or more risk factors for COVID-19 progression, and viral load at baseline was consistent with previously reported data for other anti–SARS-CoV-2 monoclonal antibodies.^39,40^ In addition, 65% of patients in this trial identified themselves as Latinx, a population that has been disproportionately affected by COVID-19 and historically underrepresented in clinical trials.^41-44^

This trial has several limitations. Given the profound efficacy of sotrovimab, a small number of events for the primary and clinical secondary outcomes were reported in patients who were randomized to sotrovimab. As a result, it is challenging to determine the patient or disease characteristics associated with COVID-19 progression in sotrovimab-treated patients. Also notable is that the moderate size of the safety population limits the ability to detect rare AEs. However, based on the development of sotrovimab from a human antibody engineered to target a viral epitope, rare AEs are not expected. Finally, the study enrolled patients over approximately 6 months, representing a finite period of the pandemic. As a result, the complete picture of viral sequencing and clinical experience with sotrovimab for variants of concern is unknown. Despite this, sotrovimab targets a viral epitope that does not overlap with mutations observed in current variants of concern, and thus it is hypothesized that sotrovimab will remain effective against these variants.^19,45^

## Conclusions

Treatment with sotrovimab reduced the progression of COVID-19 in high-risk patients with mild to moderate disease. The incidence of hospitalizations and need for supplemental oxygen was reduced with sotrovimab vs placebo. In addition, sotrovimab improved patient-reported symptomatology. This SARS-CoV-2 neutralizing antibody is an effective and well-tolerated therapy to treat early cases of COVID-19 and improve disease prognosis.

## Supporting information

CONSORT Checklist

## Data Availability

Data will serve as a basis for a regulatory filing and cannot be shared at this time.

## Author Contributions

Concept and design: AP, EA, WWY, MA

Acquisition, analysis, and/or interpretation of the data: AG, YG-R, EJ, MCC, JM, DRF, ES, JS, HZ, NS, ALC, SP, JES, DA, AP, EA, WWY, CB, MA, AES

Drafting of the manuscript: AG, YG-R, EJ, MCC, JM, DRF, ES, JS, HZ, NS, ALC, SP, JES, DA, AP, EA, WWY, CB, MA, AES

Critical revision of the manuscript for important intellectual content: AG, YG-R, EJ, MCC, JM, DRF, ES, JS, HZ, NS, ALC, SP, JES, DA, AP, EA, WWY, CB, MA, AES

Statistical analysis: HZ, NS Supervision: EA, WWY, MA

## Additional Contributions

The authors thank Courtney St. Amour, PhD, of Cello Health Communications/SciFluent, for medical writing support, which was funded by Vir Biotechnology, Inc., and GlaxoSmithKline, and Krystyna Grycz, Jordan Clark, and Minnie Kuo, of Vir Biotechnology, Inc., for clinical operations support.

