## supplementary appendix for "Effect of the Neutralizing SARS-CoV-2 Antibody Sotrovimab in Preventing Progression of COVID-19: A Randomized Clinical Trial"

**Supplement 1**

**eMethods**

**eTable 1. Primary Reasons for Hospitalizations of More Than 24 Hours or Death (ITT Population)**

**eFigure 1. Respiratory Status Over Time Excluding Room Air^a,b^**

**eMethods**

*InFLUenza Patient-Reported Outcome (FLU-PRO) adapted for COVID-19 (FLU-PRO Plus)*

The FLU-PRO is a validated, 32-item questionnaire that quantifies the presence and severity of symptoms of influenza.^1,2^ The questions span 6 body systems (nose, throat, eyes, chest/respiratory, gastrointestinal, and body/systemic) and are scored on a 5-point ordinal scale ranging from 0 (symptom free) to 4 (very severe symptoms). Responses to each item are averaged for the total score.

The FLU-PRO was adapted for COVID-19 as the FLU-PRO Plus with the addition of new items for smell and taste. Patients completed the questionnaire daily through day 21 and at day 29, week 8, and week 12.

**eTable 1. Primary Reasons for Hospitalizations of More Than 24 Hours or Death (ITT Population)^a^**

| **Treatment group** | **Patient** | **Age range, y** | **Primary reason for hospitalization or death** | **Hospitalization day** | **Hospitalization duration, days** | **ICU admission** | **Invasive mechanical ventilation** | **Fatal** |
| --- | --- | --- | --- | --- | --- | --- | --- | --- |
| Sotrovimab | 1 | 96-100 | COVID-19 pneumonia | 19 | 14 | N | N | N |
| Sotrovimab | 2 | 61-65 | Small intestinal obstruction | 22 | 3 | N | N | N |
| Sotrovimab | 3 | 51-55 | Diabetic foot ulcer | 19 | 7 | N | N | N |
| Sotrovimab | 4 | 71-75 | COVID-19 pneumonia | 2 | 16 | N | N | N |
| Sotrovimab | 5 | 56-60 | Non-small cell lung cancer | 13 | 11 | N | N | N |
| Sotrovimab | 6 | 61-65 | COVID-19 pneumonia | 2 | 5 | N | N | N |
| Placebo | 7 | 66-70 | COVID-19 pneumonia | 6 | 7 | N | N | N |
| Placebo | 8 | 36-40 | COVID-19 pneumonia | 9 | 5 | N | N | N |
| Placebo | 9 | 81-85 | Acute respiratory failure | 7 | 6 | N | N | N |
| Placebo | 10 | 81-85 | COVID pneumonia, hypovolemia | 8 | 2 | N | N | N |
| Placebo | 11 | 46-50 | COVID-19 pneumonia | 4 | 8 | Y | N | N |
| Placebo | 12 | 66-70 | Respiratory distress | 12 | 17 | Y | Y | N |
| Placebo | 13 | 66-70 | Pneumonia | 5 | 18 | Y | N | Y |
| Placebo | 14 | 71-75 | COVID-19 pneumonia | 10 | 28 | Y | Y | Y |
| Placebo | 15 | 56-60 | COVID-19 pneumonia | 2 | 31 | Y | N | N |
| Placebo | 16 | 71-75 | COVID-19 | 4 | 6 | Y | N | N |
| Placebo | 17 | 56-60 | COVID-19 pneumonia | 5 | 59 | Y | Y | N |
| Placebo | 18 | 66-70 | COVID-19 pneumonia | 5 | 9 | Y | N | N |
| Placebo | 19 | 71-75 | Respiratory failure, cardio respiratory arrest | 14 | 23 | Y | Y | Y |
| Placebo | 20 | 76-80 | COVID-19 pneumonia | N/A | *Died at home* | N | N | Y |
| Placebo | 21 | 51-55 | COVID-19 pneumonia | 4 | 6 | N | N | N |
| Placebo | 22 | 46-50 | COVID-19 pneumonia | 4 | 20 | N | N | N |
| Placebo | 23 | 61-65 | Dehydration | 6 | 7 | N | N | N |
| Placebo | 24 | 51-55 | COVID-19 pneumonia | 5 | 4 | N | N | N |
| Placebo | 25 | 56-60 | Pneumonia | 6 | 4 | N | N | N |
| Placebo | 26 | 61-65 | Pneumonia | 12 | 4 | N | N | N |
| Placebo | 27 | 66-70 | COVID-19 pneumonia | 7 | 4 | N | N | N |
| Placebo | 28 | 51-55 | Pulmonary embolism | 7 | 3 | N | N | N |
| Placebo | 29 | 56-60 | COVID-19 pneumonia | 10 | 5 | N | N | N |
| Placebo | 30 | 66-70 | COVID-19 pneumonia | 5 | 4 | N | N | N |
| Placebo | 31 | 36-40 | COVID-19 pneumonia | 6 | 8 | N | N | N |
| Placebo | 32 | 61-65 | COVID-19 pneumonia | 7 | 11 | N | N | N |
| Placebo | 33 | 71-75 | COVID-19 | 5 | 9 | N | N | N |
| Placebo | 34 | 51-55 | COVID-19 pneumonia | 7 | 5 | N | N | N |
| Placebo | 35 | 61-65 | COVID-19 pneumonia | 6 | 5 | N | N | N |

^a^Includes 4 male and 2 female patients in the sotrovimab group and 16 male and 13 female patients in the placebo group.

ITT, intent-to-treat; ICU, intensive care unit; N/A, not applicable.

**eFigure 1. Respiratory Status Over Time Excluding Room Air^a,b^**

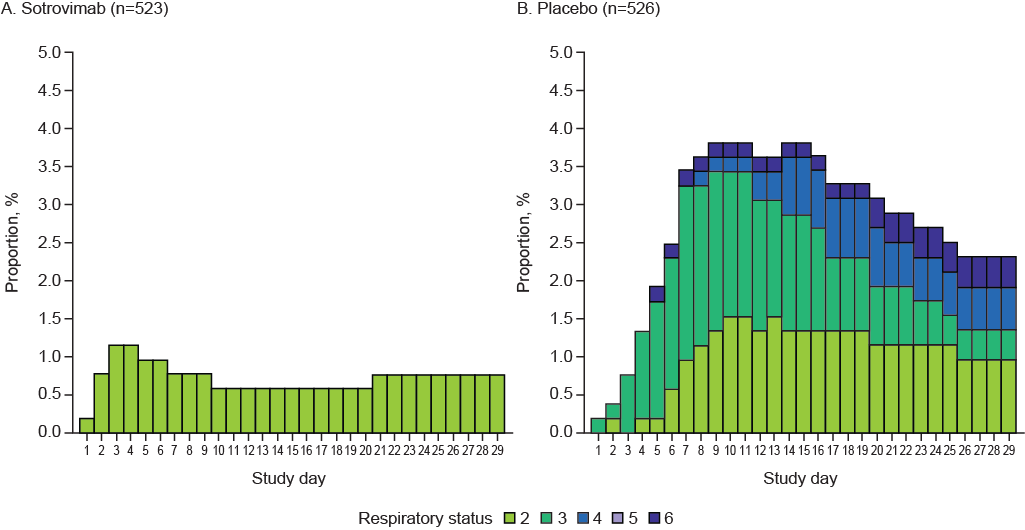

^a^Percentages are based on the number of patients observed on each study day.

^b^Respiratory support categories defined as: 2, low flow nasal cannula/face mask; 3, nonrebreather mask or high-flow nasal cannula/noninvasive ventilation; 4, mechanical ventilation/extracorporeal membrane oxygenation; 5, other; 6, death.
